# High viral SARS-CoV-2 load in placenta of patients with hypertensive disorders after COVID-19 during pregnancy

**DOI:** 10.1101/2021.09.07.21261607

**Authors:** M Fabre, P Calvo, S Ruiz-Martinez, M Peran, D Oros, A Medel Martinez, M Strunk, R Benito, J Schoorlemmer, C Paules

**Affiliations:** Instituto de Investigación Sanitario de Aragón (IIS Aragon) Biochemistry Department, Hospital Clínico Universitario Lozano Blesa Zaragoza, Spain; Placental pathophysiology & fetal programming research group, B46_20R & GIIS-028 del IISA; Instituto de Investigación Sanitario de Aragón (IIS Aragon), Obstetrics Department, Hospital Clínico Universitario Lozano Blesa Zaragoza, Spain; Red de Salud Materno Infantil y del Desarrollo (SAMID), RETICS. Instituto de Salud Carlos III (ISCIII), Subdirección General de Evaluación y Fomento de la Investigación y Fondo Europeo de Desarrollo Regional (FEDER) Ref: RD16/0022/0013; Laboratorio Satélite, Instituto Aragonés de Ciencias de la Salud (IACS), Centro de Investigación Biomédica de Aragón (CIBA), Zaragoza, Spain; Sequencing and Functional Genomics, Instituto Aragonés de Ciencias de la Salud (IACS), Centro de Investigación Biomédica de Aragón (CIBA), Zaragoza, Spain; Microbiology Department, Hospital Clínico Universitario Lozano Blesa, Zaragoza, Spain. Universidad de Zaragoza. IIS Aragon; Instituto Aragonés de Ciencias de la Salud (IACS), Zaragoza, Spain; ARAID Foundation, Zaragoza, Spain

**Keywords:** SARS-CoV-2, placenta, hypertensive disorders of pregnancy, COVID-19, placental viral load

## Abstract

**Introduction:** Studies described an increased frequency of hypertensive disorders of pregnancy after a COVID-19 episode. There is limited evidence about SARS-CoV-2 viral load in placenta. This study aimed to investigate the relationship between SARS-CoV-2 viral load in placenta and clinical development of HDP after COVID-19 throughout different periods of gestation.

**Methods:** This was a case-control study in women with and without gestational hypertensive disorders (HDP) after SARS-CoV-2 infection diagnosed by RT-PCR during pregnancy. Patients were matched by gestational age at the moment of COVID-19 diagnosis. We performed an analysis of SARS-CoV-2 RNA levels in placenta.

**Results:** A total of 28 women were enrolled. Sixteen patients were diagnosed with COVID-19 during the third trimester and the remaining twelve patients in the others trimesters. Ten placentas (35.7%) were positive for SARS-CoV-2, nine of them (90%) belonged to the HDP group versus one (10%) in control group (p=0.009). Those cases with the highest loads of viral RNA developed severe-preeclampsia.

**Conclusion:** The presence of SARS-CoV-2 was more frequent in placentas of patients with HDP after COVID-19. There seems to be a relationship between high viral load in the placenta and the development of hypertensive disorders. We found SARS-CoV-2 viral load in placenta after birth in mothers infected at the first half of pregnancy, but with negative nasopharyngeal RT-PCR at delivery. Our data suggest that SARS-CoV-2 infection during pregnancy could trigger gestational hypertensive disorders through placenta-related mechanisms.

## INTRODUCTION

Several international studies have recently suggested an increase in preeclampsia and other hypertensive disorders of pregnancy (HDP) associated with SARS-CoV-2 infection during pregnancy [1,2,3,4] with classic signs and symptoms related to preeclampsia, such as hypertension, proteinuria, thrombocytopenia, elevated liver enzymes [5,6]. Preeclampsia (PE) is a pregnancy-specific multisystem disorder characterized by endothelial dysfunction that leads to the release of soluble factors into de maternal circulation responsible for the multi-organ injury [7]. These so-called preeclampsia-like syndromes are in some way clinical manifestations of maternal endothelial dysfunction.

The clinical manifestations of the SARS-CoV-2 infection support a key role for endothelial dysfunction in the pathobiology of this condition. SARS-CoV-2 infection induces an endotheliitis in different organs as a direct consequence of viral infection and the host inflammatory response [8,9]. Angiotensin-converting enzyme 2 (ACE2) is the major cellular-entry receptor for SARS-CoV-2 virus [10]. ACE2 is present prominently in the alveoli, but also in endothelial cells, which can be directly infected by the virus [11]. Besides, cytokine release syndrome can also drive endothelial damage independently [12]. Consequently, a distinctive feature of SARS-CoV-2 infection is vascular harm, with severe endothelial injury, widespread thrombosis and microangiopathy, in response to endothelial damage. Therefore, endothelial dysfunction seems to be the pathophysiological substrate for severe COVID-19 complications.

It is at present unknown how COVID-19 is associated with increased frequency of HDP (and especially PE) and whether similar mechanisms operate. COVID-19 associated PE-like disease may be an indirect result of maternal disease or more directly related to the presence of SARS-CoV-2 in the placenta. While initial studies had suggested that SARS-CoV-2 invades the placenta in some cases [13, 14, 15], posterior data suggest that is not very common [16, 17, 18, 19]. Consistent with a low frequency presence of SARS-CoV-2 in the placenta [22], vertical transmission is rather infrequent [21]: a systematic review reported a rate of 3.2% vertical transmission in mothers who tested positive for SARS-CoV-2 (diagnosed by RT-PCR) during the third trimester of pregnancy [22]. A recent study of placentas taken from COVID-19 positive mothers [23] describes a low-level presence of RNA in about half of placentas. No substantial histopathological, maternal, or neonatal outcome features were detected associated with this low level of viral RNA. However, a high SARS-CoV-2 RNA level was detected in only one patient, who exhibited severe placental injury in the form of extensive fibrin deposition, necrosis of the syncytiotrophoblast layer of the villi and apoptosis. This observation fits the hypothesis that only limited placental pathology is associated with COVID-19, except for cases with very high SARS CoV2 RNA levels, the latter indicative of high viral load.

Considering the increase in hypertensive disorders in COVID-19 patients, this study aimed to investigate the relationship between SARS-CoV-2 viral load in placental tissue at birth and the development of preeclampsia or gestational hypertension after COVID-19 throughout different periods of gestation. The clinical history of three cases of preeclampsia, with positive placental PCR tests for SARS-CoV-2 during each of the three semesters of pregnancy, is described in detail as an example.

## Materials and Methods

We conducted a case-control study in women diagnosed with COVID-19 during pregnancy with and without gestational hypertensive disorders. We perform an analysis of SARS-CoV-2 RNA levels in placental tissue and SARS-CoV-2 antibodies in umbilical cord. Patients were recruited at the moment of delivery between May 2020 and February 2021 at a tertiary University Center. Cases were defined as women who tested positive for SARS-CoV-2 during pregnancy and were diagnosed with gestational hypertensive disorders. Controls, defined as SARS-CoV-2 positive women without HDP during pregnancy, were paired by gestational age at the moment of SARS-CoV-2 diagnosis.

SARS-CoV-2 infection was diagnosed based on positive RT-PCR test for SARS-CoV-2 from nasopharyngeal swabs. RT-PCR test kits from different companies were used: Viasure (CerTest Biotec, Zaragoza, Spain), M2000 SARS-CoV-2 Assay (Abbott RealTime SARS-CoV-2 Assay, Abbott Molecular, Abbott Park, IL, USA), TaqPath COVID-19 (Thermo Fisher Scientific, USA-FDA) and Alinity SARS-CoV-2 (Abbott Alinity, Abbott Molecular, Abbott Park, IL, USA). Information on these test kits are listed in Supplementary Table1. As suggested by the manufacturer for nasopharyngeal specimens, cycle threshold (CT) values below 37 were taken as positive. Besides, all women who were hospitalized for delivery were screened for SARS-CoV-2 infection using RT-PCR on nasopharingeal swabs shortly before giving birth. However, in the case of pregnant women who had already overcome the infection, PCR-RT was only performed around delivery if the latest negative PCR test was more than 3 months ago. As a result, four patients (3, 4, 15 and 16) were not tested for SARS-CoV-2 infection at the time of delivery. Prophylactic treatment with low molecular weight heparin for 14 days was applied to COVID-19 women diagnosed by RT-PCR [24]. COVID-19 has been divided into three types [25]: asymptomatic infection refers no clinical symptoms or signs; mild infection refers symptoms such as fever, cough, headache, anosmia and asthenia; and severe infection refers dyspnea, hypoxemia accompanied by chest imaging compatible with pneumonia and respiratory infection.

Placental tissue samples were taken at the moment of delivery, taking villous tissue while carefully trying to avoid the overlying maternal tissue. Placental samples, approximately 1 cm^3^ in size, were placed into tubes containing 1 ml of preservative solution (RNAlater Fisher Scientific) at 4°C. After 24h, excess RNAlater was removed, and the samples were stored at -80°C. Tissue was homogenized, RNA was extracted, treated with DNase I, repurified and resuspended as previously described [26]. Concentrations were measured using Nanodrop, 100 ng of RNA was used in downstream PCR analysis. RT-PCR for SARS-CoV-2 in placenta tissue samples was performed using the TaqPath COVID-19 CE-IVD RT-PCR kit (Catalog Number A48067; Life Technologies Europe, Bleiswijk, The Netherlands) on a QuantStudio5 (Applied Biosystems; ThermoFisher Scientific) apparatus. The limit of detection of this kit is listed as 10 GCEs (Genomic Copy Equivalents). As this assay targets N, ORF1ab and S genes, only samples with three positive targets were considered as SARS-CoV-2 positive. SARS-CoV-2 tissue load in placenta is referred to in the manuscript as viral load and is expressed as CT values of RT-PCR.

We collected umbilical cord plasma in EDTA and samples were centrifuged. Serological assays were determined on the same day by chemiluminescent microparticle immunoassays for quantitative detection of SARS-CoV-2 IgG. The SARS-CoV-2 IgG assay is designed to detect IgG antibodies to the nucleocapsid protein of SARS-CoV-2. (IgG; SARS-CoV-2 IgG Assay, Abbott Laboratories Ireland, Dublin, Ireland) and IgM+IgA antibodies (IgM+IgA; COVID-19 VIRCLIA IgM+IgA, Vircell Microbiologists, Granada, Spain).

Gestational hypertensive disorders were divided into four categories, defined according to the criteria proposed by American college of Obstetrics and Gynaecologists [27]: preeclampsia, chronic hypertension, chronic hypertension with superimposed preeclampsia and gestational hypertension. First trimester risk of early-onset preeclampsia was retrospectively calculated according to maternal characteristics, obstetric history, maternal blood pressure, maternal serum pregnancy-associated plasma protein-A and uterine artery pulsatility index, using SsdwLab6 version 6.1 package (SBP Soft 2007 S.L.) [28].

Clinical characteristics, laboratory results, maternal and neonatal outcomes were collected from medical records. All patients provided written informed consent. The study was approved by the Research Ethics Committee of the Community of Aragon (C.I. PI21/155 and COL21/000) and all patients provided written informed consent.

Statistical analysis was performed using SPSS 22.0. Categorical variables were presented as frequencies or percentages. Continuous variables were presented using mean ± standard deviation, median, or range. For continuous variables, Shapiro Wilk tests of normality were used to evaluate the distributions. Data were analysed using Student t test or Mann–Whitney U test. Statistical significance was considered p < 0.05.

## Results

Fourteen women in the case group (COVID-19 positive and diagnosis of HDP) and fourteen women in the control group, matched for gestational age at the time of COVID-19 diagnosis, were included. Out of those 14 cases, 9 cases (64.3%) had preeclampsia and five cases (35.7%) had gestational hypertension. Demographic and clinical data, as well as perinatal and neonatal outcomes are shown in table 1. There were no significant differences in maternal age, maternal weight, risk of preeclampsia at 1º trimester, nulliparity rate and diabetes mellitus between the two groups. The vast majority of patients, eight pairs, represent pregnant women who were diagnosed with COVID-19 during the third trimester, whereas five pairs in the second trimester and only one pair in the first trimester. Mean gestational age at delivery was 268.1 days in the case group and 278.9 days in the control group (p=0.005). Women with HPD also exhibited lower birthweight compared with the control group (2852.5g SD 467.0 versus 3282.9g SD: 435.9; p=0.005). There were no significant differences for small for gestational age, umbilical arteria ph<7.1. Induced labor and cesarean was significantly more frequent in the HDP group compared to controls (p= 0.003 and p=0.015, respectively).

**Table 1.** Demographic data and obstetrical and SARS-CoV-2 infection characteristics.

|  | CASE GROUP (n=14) | CONTROL GROUP (n=14) | p |
| --- | --- | --- | --- |
| **Demographic maternal data** |  |  |  |
| Maternal age (SD), years | 31.93 (6.6) | 30.36 (5.8) | 0.506 |
| Maternal weight (SD), kg | 69.32 (10.5) | 68.57 (13.2) | 0.868 |
| Caucasian (%) | 9 (64.3) | 4 (28.6) | 0.063 |
| 1<sup>o</sup> trimester high risk for Preeclampsia (%) | 5 (35.7) | 3 (21.4) | 0.194 |
| Diabetes mellitus (%) | 0 (0) | 1 (7.14) | 0.317 |
| Nullipara (%) | 4 (28.6) | 7 (50) | 0.317 |
| **Maternal and neonatal outcome at delivery** |  |  |  |
| Gestational age at birth mean (SD), days | 268.1 (10.8) | 278.9 (7.8) | 0.005 |
| Birth weight, mean (SD), kg | 2852.5 (467.0) | 3282.9 (435.9) | 0.018 |
| SGA (<10th centile) (%) | 4 (28.6) | 1 (7.2) | 0.146 |
| Induced labor (%) | 12 (85.1) | 4 (28.6) | 0.003 |
| Cesarean delivery (%) | 5 (35.7) | 0 (0) | 0.015 |
| pH<7.1 (%) | 2 (14.3) | 0(0) | 0.541 |
| Preterm birth (<37 wk gestation) (%) | 3 (21.4) | 0(0) | 0.072 |
| **Description of SARS-CoV-2** |  |  |  |
| Trimester of SARS-CoV-2 infection |  |  |  |
| 1<sup>o</sup> (%) | 1 (7.2) | 1 (7.2) | 1 |
| 2<sup>o</sup> (%) | 5 (35.7) | 5 (35.7) |  |
| 3<sup>o</sup> (%) | 8 (57.1) | 8 (57.1) |  |
| COVID-19 Symptoms |  |  |  |
| No | 6 (42.9) | 7(50) | 0.341 |
| Mild | 6 (42.9) | 7 (50) |  |
| Severe | 2 (14.2) | 0 (0) |  |
| Interval from diagnosis of SARS-CoV-2 to delivery (SD), d | 61.86 (54.79) | 72.64 (57.7) | 0.616 |
| Positive SARS-CoV-2 RT-PCR Placenta tissue (CT<37) (%) | 9 (64.3) | 1 (7.2) | 0.009 |
| Neonatal IgG (%)* | 6 (42.9) | 7 (50) | 0.688 |
| Table 1. SD: standard deviation; CT: cycle threshold value RT-PCR; SGA: small for gestational age<br>*Analyzed 24 of 28. |  |  |  |

The chronology of diagnosis of HDP and SARS-CoV-2 infection of each patient is summarized in table 2. There was no significant difference in the severity of SARS-CoV-2 symptoms initial between the groups (p= 0.341). Thirteen had no symptoms, thirteen patients had mild and only two patients had severe SARS-CoV-2 infection (Table 2). No patient required admission to the intensive care unit.

**Table 2.**
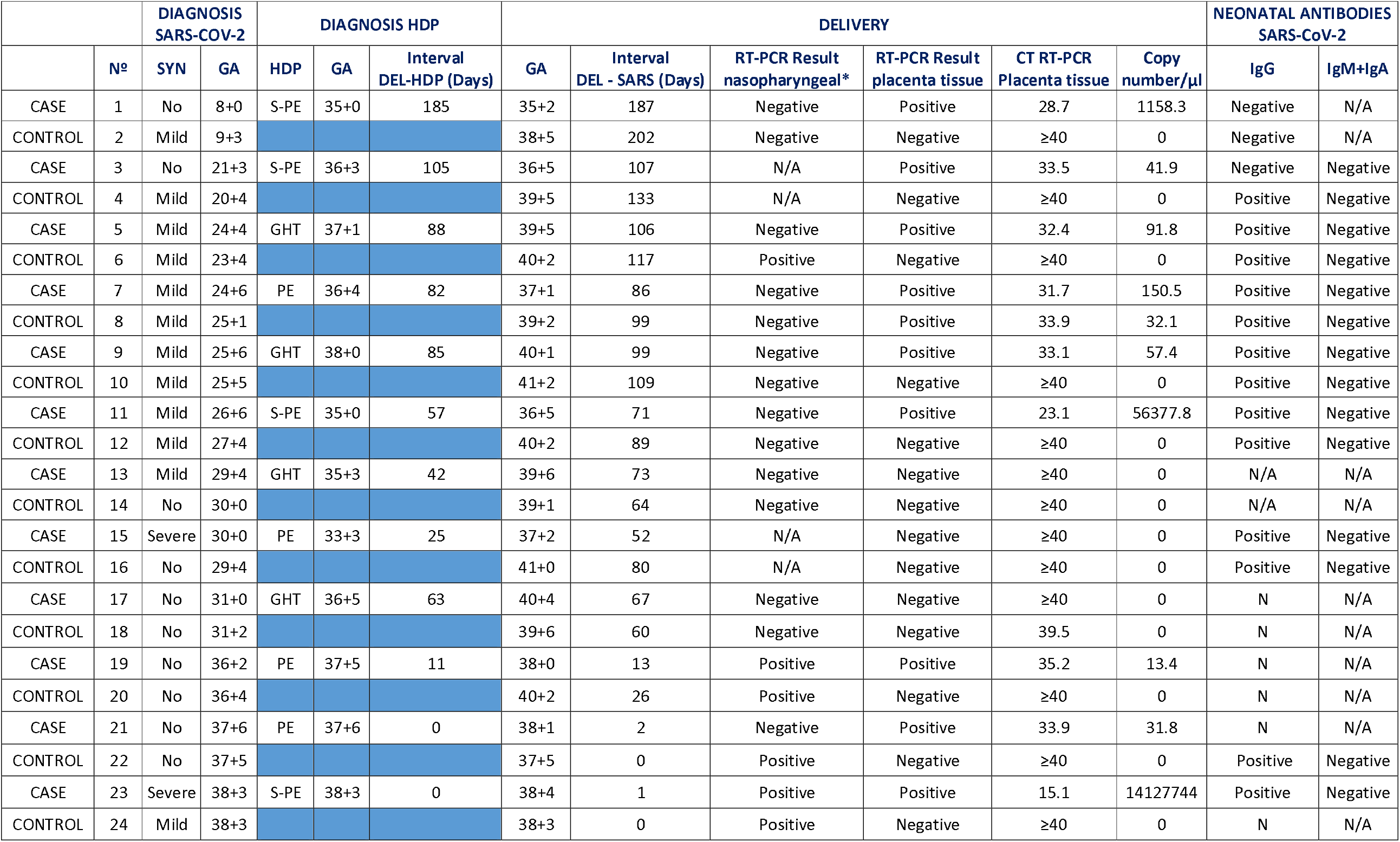
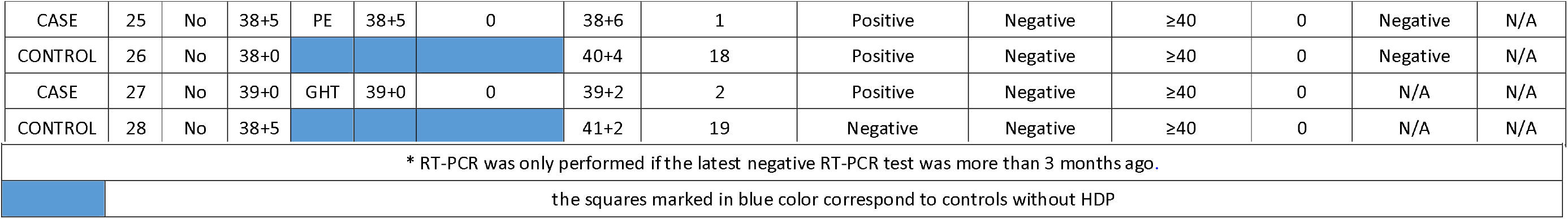
Syn: Symptoms of SARS-COV-2 infection; GA: gestational age (week + days); HDP: hypertensive disorders of pregnancy; GHT: Gestational hypertension; PE: preeclampsia; S-PE: severe preeclampsia; DELIV-PE (Days):interval from diagnosis of COVID19 to HDP diagnosis, days; DELIV-SARS (Days): interval from diagnosis of COVID19 to delivery, days; CT: cycle threshold value RT-PCR; R: result; P: positive; N: negative; N/A: not available;

Using a sensitive RT-PCR assay, ten placentas (35.7%) from SARS-CoV-2 positive women during pregnancy, tested positive for SARS-CoV-2 RNA at birth. In the case group, we found nine positive placentas (9/10, 90%) while in control group only one placenta (1/10, 10%) were positive (p=0.009) (shown in Fig.1). Moreover, 7 of nine 9 placentas (77.8%) belong to women diagnosed with preeclampsia. Focusing on positive placentas, we have three cases with high loads of viral RNA (CTs ≤ 28) (see below), and the remaining seven cases with low loads of viral RNA (median CT of 30.6; Interquartile range 28.7-33.9) (Table 2).

**Figure 1.**
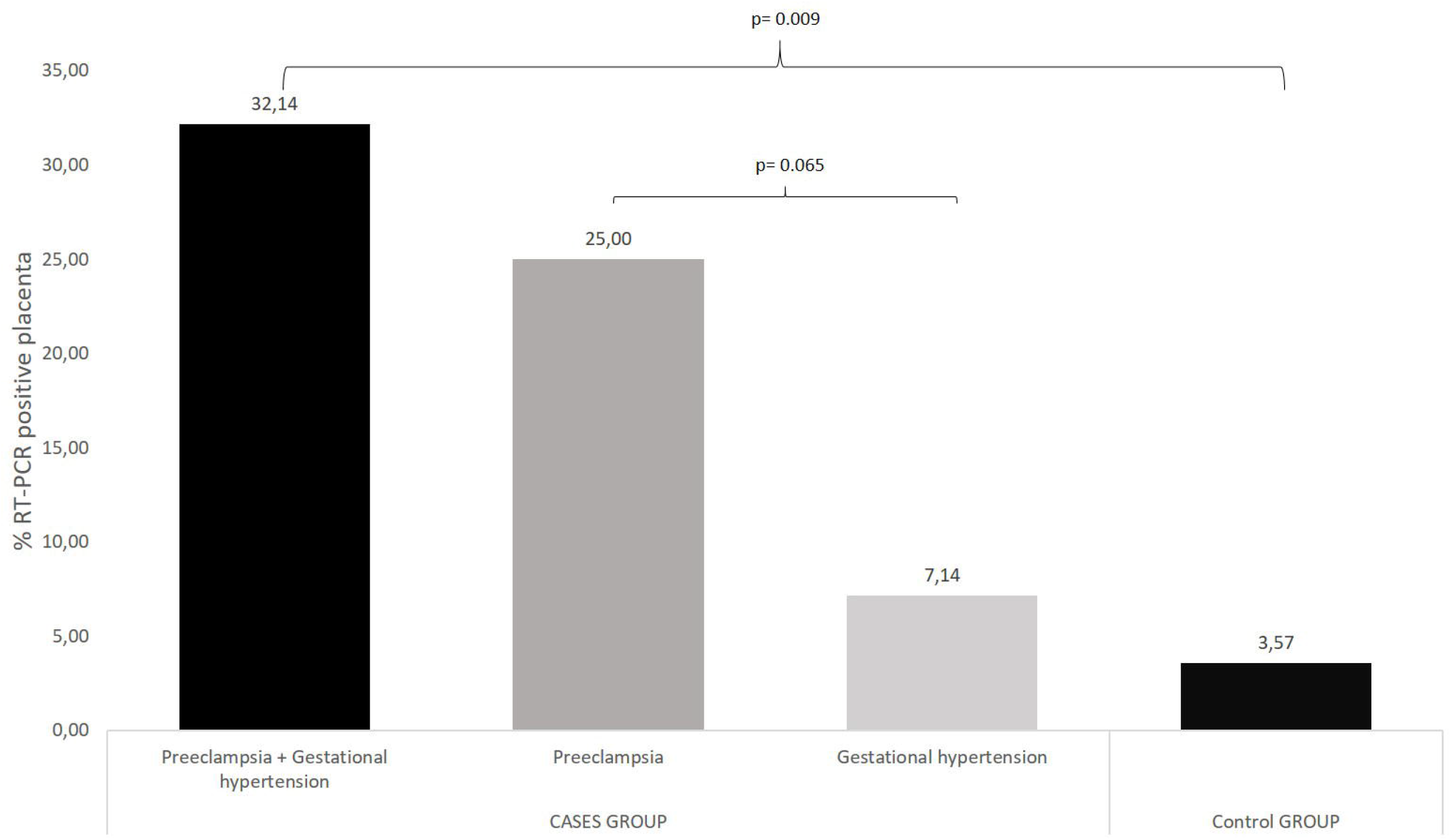
% RT-PCR positive placenta: cases group vs control group. Ten placentas (35.7% = 3.57% + 32.14%) tested positive for SARS-CoV-2 RNA. Nine positive placentas in the cases group (9/28, 32.14%) while in control group only one placenta were positive (1/28, 3.57%). Seven (7/28, 25%) belong to women diagnosed with preeclampsia vs two of nine belong to women diagnosed with gestational hypertension (2/28, 7.14%).

While only six pairs of pregnancies with positive SARS-CoV-2 tests during the first or second trimester (GA ≤ 27 weeks) were analyzed (Table 2), several remarkable results deserve mentioning. First, out of the six cases with HDP, six out of six had positive placental SARS-CoV-2 test at birth. In contrast, among the corresponding six control cases without HDP, only one tested positive for placental SARS-CoV-2 (Table 2).It is possible therefore, that the continued presence of SARS-CoV-2 in the placenta promotes HDP or vice versa. Moreover, out of ten pregnancies for which data are available with these early SARS-CoV-2 infections, only one mother remained positive at delivery (Nº 6), while seven placentas tested positive. This latter data suggests that infection with SARS-CoV-2 during early pregnancy may commonly result in persisting placental presence of SARS-CoV-2.

Finally, antibody quantification was performed for 87.5% neonates with umbilical cord blood samples. Thirteen infants tested positive for IgG on the first 10 days of life, and none tested positive for IgM+IgA. There was no significant difference between the groups in the number of neonates that tested positive for SARS-CoV-2 IgG (p= 0.688).

Among the placentas that tested positive, three cases with high loads of viral RNA (Cts = 15; 23; 28, respectively) represented cases of severe preeclampsia (Table 2). Case number 1 (shown in Fig.2) was a woman with no medical history of interest. The estimated risk of early-onset preeclampsia in the first trimester was 1/122. She presented positive SARS-CoV-2 RT-PCR (CT: 18.7) at week 8 of gestation and infection courses asymptomatic. Two weeks later, she tested negative for SARS-CoV-2 by RT-PCR. At 35 week of gestation, she debuted with severe preeclampsia, which required admission to the hospital and treatment with intravenous labetalol and magnesium sulfate. RT-PCR for SARS-CoV-2 infection was performed at the time of admission, with a negative result. Termination of pregnancy was in week 35 + 2 for this reason. A female of 1875 grams, APGAR test was 9 and 10 at 1 and 5 minutes, respectively, and pH 7.31 without antibodies against SARS-CoV-2 was born. Placental analysis for SARS-CoV-2 was positive, with a high viral load by RT-PCR.

**Figure 2.**
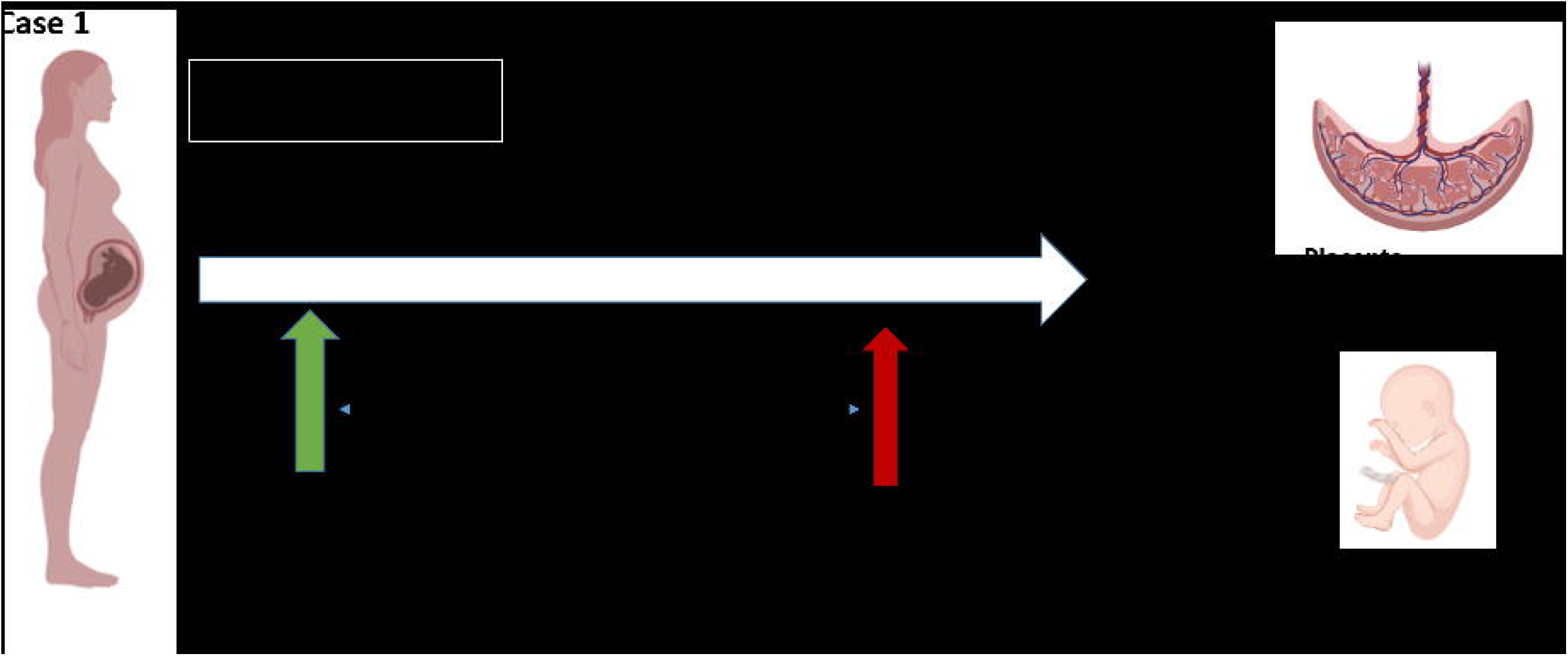
Timeline of case 1. TPAL: term, pre-term, abortion, life; LMWH: low molecular weight heparin; PAPP-A: pregnancy-associated plasma protein-A; MoM: multiple of median; BP: Mean Arterial Blood Pressure; UtA PI: uterine artery resistance pulsatility index; w: weeks; EFW: estimated fetal weight; PI UA: umbilical artery pulsatility index; PI MCA: fetal middle cerebral artery pulsatility index; IUGR: Intrauterine growth restriction; BP: blood pressure.

Case number 11 (shown in Fig.3) was a nulliparous woman with a spontaneous monochorionic diamniotic twin. The estimated risk of early-onset preeclampsia in the first trimester was 1/15. Pregnancy of normal course until week 26+5, when she debuted with fever, cough and myalgias. RT-PCR results for SARS-CoV-2 was positive (CT: 5.9). At 34+0 she tested negative for SARS-CoV-2 by RT-PCR. In week 35, the patient presented non-severe preeclampsia, well controlled with oral treatment with alfamethyldopa. At 36+5 weeks was decided an elective delivery due to the onset of severe preeclampsia, ending in an intrapartum caesarean section. The first male twin was born: 3320 grams, APGAR test was 9 and 10 at 1 and 5 minutes, respectively, and was admitted for observation for 2 days. Second twin, male, 2360 grams, with APGAR test of 4 and 8 at 1 and 5 minutes, and he was admitted for observation for 6 days. Cord blood serology was positive for IgG and negative for IgM. The placenta was clearly positive for SARS-CoV 19 by RT-PCR.

**Figure 3.**
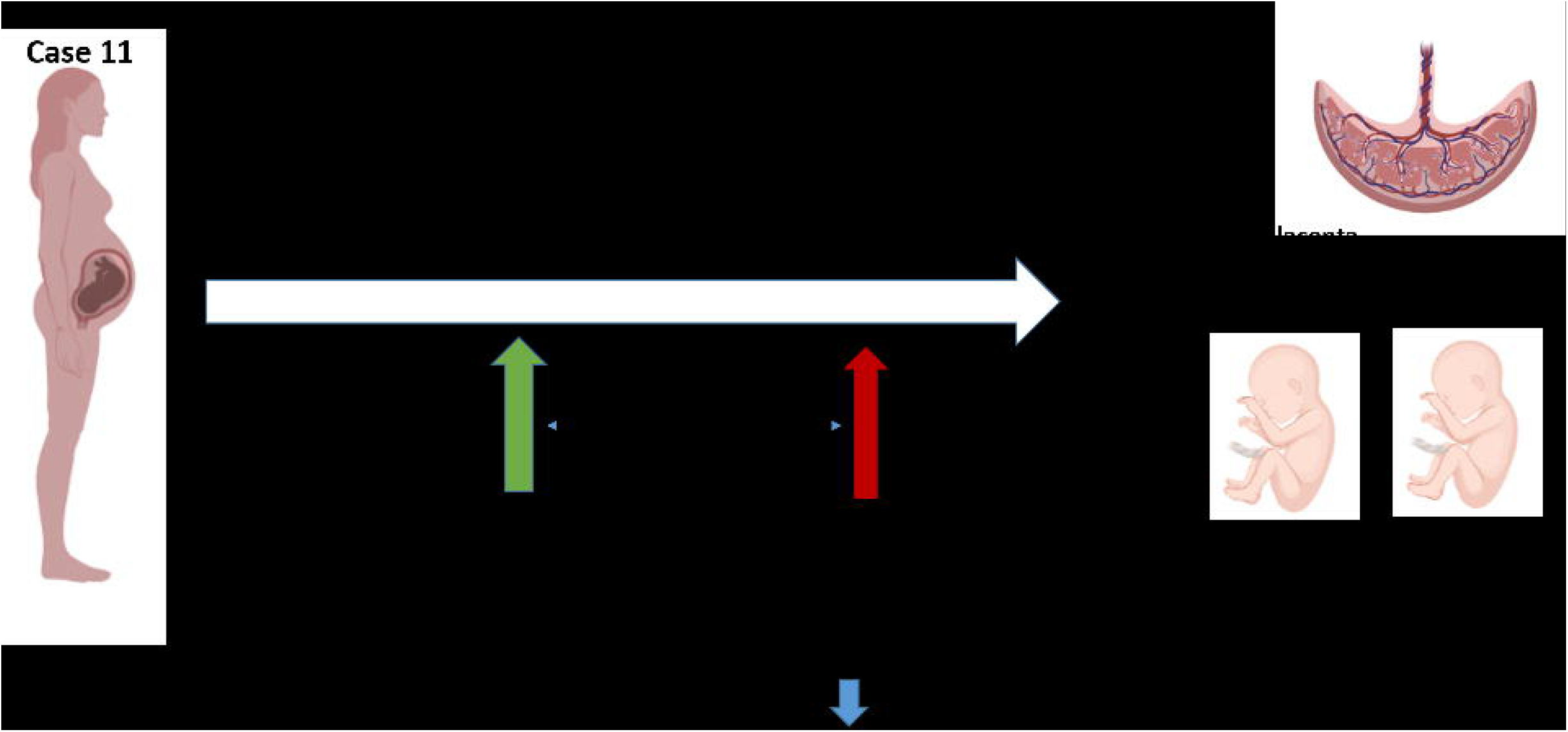
Timeline of case 11. TPAL: term, pre-term, abortion, life; LMWH: low molecular weight heparin; PAPP-A: pregnancy-associated plasma protein-A; MoM: multiple of median; BP: Mean Arterial Blood Pressure; UtA PI: uterine artery resistance pulsatility index; w: weeks; EFW: estimated fetal weight; PI UA: umbilical artery pulsatility index; PI MCA: fetal middle cerebral artery pulsatility index; IUGR: Intrauterine growth restriction; BP: blood pressure; NA: neonatal admission.

Case 23 (shown in Fig.4) was a woman with a bi-chorionic biamniotic twin pregnancy. The estimated risk of early-onset preeclampsia in the first trimester was 1/4440. Gestation of normal course until onset of fever, cough and malaise, with positive result in RT-PCR SARS-CoV-2 (CT: 20.2) at 38 +3 weeks of gestation. The patient presented blood pressure around 150/90 mmHg, with positive creatinine protein ratio and a pneumonia on chest X-ray that required treatment with low molecular weight heparin, steroids and oxygen therapy. Laboratory tests were performed in week 38+4, HELLP syndrome was detected (59000 mm3 platelets, LDH 443 U/L and GOT 43 U/L), and it was decided to elective delivery at that time by caesarean section due to HELLP and breech presentation. The birthweight of the first male twin was 2820 grams, with APGAR test 6 and 8 at 1 and 5 minutes, respectively. The birthweight of the second male twin was 2840 grams, APGAR test was 2 and 8 at 1 and 5 minutes. Both were admitted to the neonatal unit and the mother had no contact with the newborns after the cesarean section. RT-PCR SARS-CoV-2 was performed on both, positive at 3 and 2 days of life respectively, as well as SARS-CoV-2 serologies, which were positive at 19 and 16 days of life respectively. The analysis of placenta yielded a clear positive result for SARS-CoV-2 (CT 23.1) [6].

**Figure 4.**
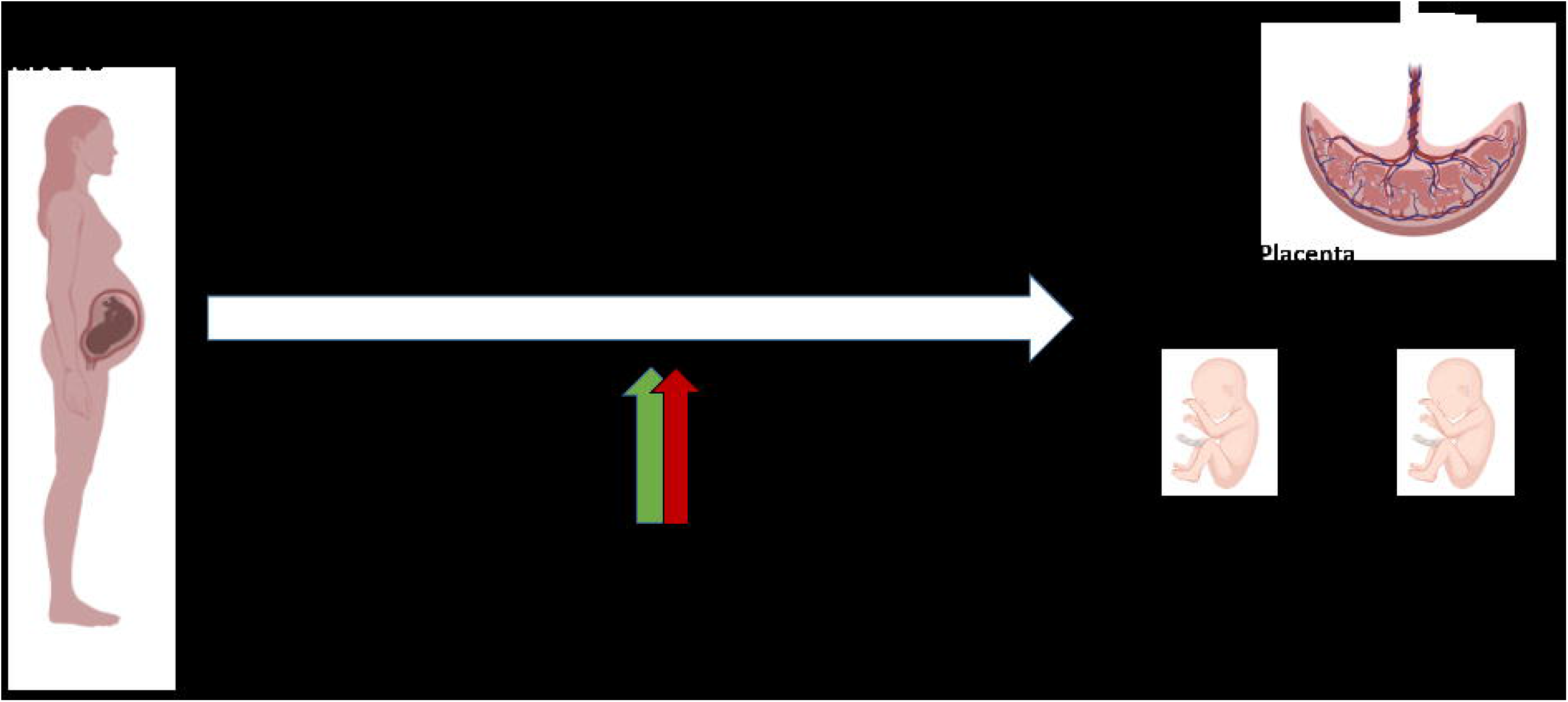
Timeline of case 23. TPAL: term, pre-term, abortion, life; LMWH: low molecular weight heparin; PAPP-A: pregnancy-associated plasma protein-A; MoM: multiple of median; BP: Mean Arterial Blood Pressure; UtA PI: uterine artery resistance pulsatility index; w: weeks; EFW: estimated fetal weight; PI UA: umbilical artery pulsatility index; PI MCA: fetal middle cerebral artery pulsatility index; IUGR: Intrauterine growth restriction; BP: blood pressure; NA: neonatal admission.

## Discussion/Conclusion

Previous viral epidemics have confirmed that pregnant women are at increased risk for severe virus infection [30, 31, 32]. SARS-COV-2 is not an exception, as pregnant patients with COVID-19 are at increased risk for severe illness [33, 34] and adverse pregnancy outcomes [8]. Therefore, women with COVID-19 may require closer control than non-infected pregnant population.

Placental histopathology at term in women suffering COVID-19 has been examined in relation to disease symptoms such as systemic inflammatory responses, hypercoagulation and intervillous thrombi [35, 36]. The presence of SARS-CoV-2 in the placenta has been studied in the context of horizontal transmission of the virus [22] with an early study reporting 7.7% positivity (2/26). However, only a very limited number of studies report data that allow a direct comparison between the placental presence of SARS-CoV-2 and HDP. An early prospective cohort study [16] describes the inability to detect SARS-CoV-2 in placenta by RNA in situ hybridization in 44 women with SARS-CoV-2 diagnosis in the third trimester. Afterwards, since the first report describing the detection of SARS-CoV-2 fetal membrane samples [37], similar reports have followed. Hosier et al [38] mentions high viral load in the placenta (CT<16) of a single woman with severe preeclampsia and SARS-CoV-2 infection during the late second trimester of pregnancy. In a posterior study, a rate of 47% SARS-CoV-2 positive placenta tissue was reported in 21 women who were diagnosed (using RT-PCR) between 35 and 40 weeks of gestation [23]. In this latter study, a woman who exhibited severe placental damage delivered a baby with neurological manifestations. Based on these combined two cases, the possibility was raised that the severity of SARS-CoV-2-mediated placental pathology might be directly related to placental SARS-CoV-2 viral load.

Following up on these studies, we further assessed the relationship between the SARS-CoV-2 viral load in placental tissue and the development of HDP in 14 HDP cases and paired controls, all diagnosed with COVID-19. Out of 28 samples tested in our study, we identified 10 placental tissue samples that tested positive for SARS-CoV-2 (35.7% of total), nine of them belonging to the hypertension disorders during pregnancy (HDP) group. We observed that, among patients with SARS-CoV-2 infection during gestation, the frequency of a positive placental SARS-CoV-2 PCR test was much higher in the HDP group compared to the non-HDP group. Furthermore, we detected that the percentage of cases with a positive placental SARS-CoV-2 RT-PCR test trend differently between PE and AHT (p= 0.065). In fact, the three most severe cases of preeclampsia (cases Nº 1, 11 and 23), showed higher placental viral load than all the others. Moreover, each of the three tested positive for SARS-CoV-2 in a different trimester of gestation. It has been argued that localization of SARS-CoV-2 to fetal cells such as the syncytiotrophoblast as opposed to maternal uterine cells requires detection by immunohistochemistry or RNA in situ hybridization [16]. We only applied RT-PCR and can therefore not exclude that the placenta samples we have analyzed do contain maternal tissue. Considering that maternal tissue was avoided when taking samples, and for the sake of simplicity, we interpret positive placental RT-PCR results as representative of viral load in villous placenta. This might indicate that in women susceptible to HDP, the presence of placental SARS-CoV-2 may contribute to the severity of the disorder. We suggest that a relationship between a higher viral load in placenta tissue and the severity of the hypertensive disorder deserves consideration.

Placental characteristics described in SARS-CoV-2 infection include maternal vascular perfusion and inflammation [35, 39]. It has been postulated that the detrimental health outcomes associated with HDP are the result of generalized endothelial and vascular dysfunction [40]. Similarly, a consequence of SARS-CoV-2 infection is endothelial injury in different organs resulting directly or indirectly from the host inflammatory response [9]. Alternatively [41], it has been proposed that direct infection of syncytioprophoblast cells in the placenta by SARS-CoV-2 may cause placental dysfunction and pregnancy complications [42]. The pathophysiological mechanisms that connect placental SARS-CoV-2 with HDP remain to be established.

The main limitation of our study is the small sample size. Further studies with a large number of patients are required to confirm the association between high SARS-CoV-2 viral load in placenta tissue and the development of HDP. Moreover, our hospital’s protocols changed as the pandemic progressed. Therefore, different methodologies have been used for viral RNA detection. As not all tests proved to be equally sensitive and specific, the use of distinct tests may have caused a discrepancy with studies published previously [13, 16, 22]. In addition, we do not have sequencing data from nasopharyngeal maternal swabs available, but all pregnant women were diagnosed before the alpha (B.1.1.7 UK) variant and delta (B.1.617.2 India) were routinely detected in our area. A final limitation to address is the fact that for each placenta we tested only a single sample. As SARS-CoV2 is not necessarily homogenously present (or absent) throughout the placenta, the identification of samples as positive or negative should be interpreted in this context.

Among the strengths of the study, we highlight the inclusion of women who tested positive for SARS-CoV-2 in each of the three trimesters of pregnancy. We demonstrate a higher viral load in placental tissue in the case group than in the control group and a higher frequency of infected placentas. Besides, as SARS-CoV-2 RT-PCR test carried out at the time of birth showed all mothers tested negative, we could rule out the possibility of acute maternal infection during delivery. Some cases we describe with high placenta SARS-CoV-2 levels, specifically patient 1 and patient 11, had negative RT-PCR results at the time of delivery. This data shows that placenta infection may persist in the absence of maternal infection. In the limited of cases we describe, this persistent viral presence is associated with the development of adverse perinatal outcomes.

In summary, this study assess the relationship between SARS-CoV-2 viral load in placental tissue and the maternal risk of developing gestational hypertensive disorders. We find that the presence of SARS-CoV-2 (although we do not formally prove viral infection) was more frequent placentas of the HDP group compared to placentas of COVID-19 mothers without HDP. We detect placental RNA representative of SARS-CoV-2 infection at high levels in a limited number of PE cases, and at lower levels in about 25% of placentas from COVID-19 positive mothers. There are different ways in which the infant of a PCR positive mother could be affected by SARS-CoV2: maternal inflammation and endothelial damage may be transmitted to the placenta in the form of cytokines which might modulate the fetal immune system [43]. Alternatively, SARS-CoV-2 infection of the placenta may directly influence the fetal environment. The presence of placental viral RNA needs to be investigated in a much larger cohort to clarify if and in what frequency if SARS-CoV-2 infection during pregnancy does trigger gestational hypertensive disorders through placenta-related mechanisms. Our work contributes to the understanding of how SARS-CoV-2 virus affects pregnancy, which includes the possibility that placental infection contributes to pregnancy complications. Better and more detailed understanding of the placental disease process may aid or could be essential for appropriate monitoring during pregnancy and providing appropriate postnatal care.

## Data Availability

All data generated or analyzed during this study are included in this article and its supplementary material files. Further enquiries can be directed to the corresponding author.

