## Supplemental table 1 for "High viral SARS-CoV-2 load in placenta of patients with hypertensive disorders after COVID-19 during pregnancy"

|  | **Nº** | **Rt-PCR result by nasopharyngeal swabs** | **CT** | **Technology** |
| --- | --- | --- | --- | --- |
| CASE | 1 | Positive | 20.2 | M2000 (Abbott) |
| CONTROL | 2 | Positive | 36.6 | M2000 (Abbott) |
| CASE | 3 | Positive | 18.2/20.1 | Viasure (CerTest) |
| CONTROL | 4 | Positive | 24.14/24.94/24.41 | TaqPath (Thermo Fisher) |
| CASE | 5 | Positive | 30.4/31.4 | Viasure (CerTest) |
| CONTROL | 6 | Positive | 18.3/19.7 | Viasure (CerTest) |
| CASE | 7 | Positive | 33.3/34.3 | Viasure (CerTest) |
| CONTROL | 8 | Positive | 11.3 | M2000 (Abbott) |
| CASE | 9 | Positive | 31.7/33.3 | Viasure (CerTest) |
| CONTROL | 10 | Positive | 15.49/14.67/14.57 | TaqPath (Thermo Fisher) |
| CASE | 11 | Positive | 5.9 | M2000 (Abbott) |
| CONTROL | 12 | Positive | 17.3/18.6 | Viasure (CerTest) |
| CASE | 13 | Positive | Antigen test | PanBio (Abbott) |
| CONTROL | 14 | Positive | 20.78 | Alinity (Abbott) |
| CASE | 15 | Positive | 14.87 | M2000 (Abbott) |
| CONTROL | 16 | Positive | 31.2 | Alinity (Abbott) |
| CASE | 17 | Positive | 15.89 | M2000 (Abbott) |
| CONTROL | 18 | Positive | 21.24 | Alinity (Abbott) |
| CASE | 19 | Positive | 34.3/33.8 | Viasure (CerTest) |
| CONTROL | 20 | Positive | 22.63 | Alinity (Abbott) |
| CASE | 21 | Positive | 0/36.6 | Viasure (CerTest) |
| CONTROL | 22 | Positive | 0/37.3 | Viasure (CerTest) |
| CASE | 23 | Positive | 21.3/18.7 | Viasure (CerTest) |
| CONTROL | 24 | Positive | 30.2 | Alinity (Abbott) |
| CASE | 25 | Positive | 0/30.5 | Viasure (CerTest) |
| CONTROL | 26 | Positive | 18.35 | Alinity (Abbott) |
| CASE | 27 | Positive | 36.9 | M2000 (Abbott) |
| CONTROL | 28 | Positive | 31.5/31.5/31.9 | TaqPath (Thermo Fisher) |
| **Supplemental 1. Diagnosis of SARS-CoV-2 by nasopharyngeal swabs** | | | | |
| CT: cycle threshold value; RT-PCR;Viasure (CerTest Biotec. Zaragoza. Spain) limit of detection 40 copies/mL and target sequence: ORF1ab and N genes; M2000 SARS-CoV-2 Assay (Abbott RealTime SARS-CoV-2 Assay. Abbott Molecular. Abbott Park. IL. USA) limit of detection 100 copies/mL and target sequence: RdRp and N genes; TaqPath COVID-19 (Thermo Fisher Scientific. USA-FDA) limit of detection 40 copies/mL and target sequence: S. N AND ORF1ab genes; Alinity SARS-CoV-2 (Abbott Alinity. Abbott Molecular. Abbott Park. IL. USA) limit of detection 40 copies/mL and target sequence: ORF1ab and N genes; Panbio™ COVID-19 Ag Rapid Test (Abbott Rapid Diagnostics Jena GmbH. Germany) Positive >2.5 ng/mL SARS-CoV-2 | | | | |
